# Presence of SARS-CoV-2 in food surfaces and public space surfaces in 3 districts of Lima, Peru

**DOI:** 10.1101/2021.11.15.21265879

**Authors:** Katherine Alvis-Chirinos, Yolanda Angulo-Bazán, Oscar Escalante-Maldonado, Duilio Fuentes, Miryam Guillermina Palomino Rodriguez, Elena Gonzales-Achuy, Henry Mormontoy, Paul Hinojosa, Lucio Huamán-Espino, Juan Pablo Aparco

## Abstract

**Objective:** The goal of this study is to determine the presence of SARS-CoV-2 in food surfaces and public space surfaces in 3 districts of Lima, Peru.

**Material and methods:** Cross-sectional descriptive study, carried out in the districts of San Juan de Lurigancho, San Martin de Porres and Villa el Salvador. Surfaces that were exposed to the greatest user manipulation were selected, samples were swabbed for 4 weeks and transported to the laboratory to determine the presence of the virus.

**Results:** 1095 inert surface samples and 960 food surface samples were evaluated for the identification of SARS-CoV-2 by the RT-PCR molecular test, whereby only one sample from an ATM (Automated Teller Machine) was positive.

**Conclusions:** Most of the inert and food surfaces evaluated did not show the presence of SARS-CoV-2 during the time of sample collection. Despite the negative results, the frequency of disinfection measures should be maintained and increased, especially on inert high-contact surfaces, and hygiene measures on food should be continue.

## Introduction

An epidemic outbreak of pneumonia of unknown origin began in Wuhan-China in December 2019, which quickly spread throughout the world. The World Health Organization (WHO) named this disease COVID-19 and declared it a global pandemic in March 2020. After a year of pandemic, around 123 million confirmed cases and 2 million deaths have been reported (1). In addition to important socio-economic problems (2).

In Peru, the first case of COVID-19 was identified on March 6, 2020 (3) and the first two deaths occurred thirteen days later. As of March 2021, there are about a 1.5 million confirmed cases and 50,000 deaths (4). The Peruvian government has decreed continually a state of national emergency. The right to freedom of movement and productive activities have been limited in our country, emphasizing social isolation in order to reduce the transmission of SARS-CoV-2 (5).

COVID 19 is caused by SARS-CoV-2, an RNA virus belonging to the Orthocoronavirinae family, which also includes other agents that cause epidemics such as the Middle East Respiratory Syndrome or Acute Respiratory Syndrome Severe (SARS-CoV) (6). One of the most important characteristics of SARS-CoV-2 is its transmissibility, with basic reproduction numbers (R0), defined as the average number of new cases generated by a case (7), with a mean of 2.2, but which can range from 1.4 to 6.5 (8). This not only justifies measures of social isolation, but also requires research on other possible forms of transmission that do not involve close contact between people.

The WHO has suggested that the transmission of this virus can occur indirectly by contact with surfaces in the immediate environment or with objects used on the infected person (9). Various studies confirm that SARS-CoV-2 can remain in different types of materials from two hours to nine days (10). Although it is considered that the respiratory tract is the main form of contagion, due to the close contact that one has between people; it is postulated that the virus could remain active on surfaces and that under this route it would cause contagion by direct contact with infected objects (11). In this regard, the WHO states that viral particles are too heavy to be transported through the air and land on objects (including food) and surfaces that surround the infected person (12). Thus, viable SARS-CoV-2 virus and / or RNA detected by RT-PCR can be found on these surfaces for periods ranging from hours to days, depending on the environment (including temperature and humidity) and the type of surface.

Previous studies have reported virus transmission through food, when the mode of spread is direct (13). However, the European Food Safety Agency (EFSA) as well as the WHO have declared that food are not agents of transmission of SARS-CoV-2 (14). Non the less, the EFSA claim is based on studies related to viruses such as SARS-CoV-1. So far, studies using SARS-CoV-2 has not been directly conducted. It is important to mention the Europe has an industrialized food production and distribution, where food handling processes are mechanized, reducing the opportunity for human intervention. In our country the conditions are different, the limited regulation allows that the food is exposed to high manipulation.

There is little information on the actual viral load that can be found on inanimate surfaces and food surfaces, especially those that are frequently touched by people (15). That is why the WHO already recommends ensuring adequate disinfection and cleaning of these surfaces. Most of the studies that have addressed the coronavirus burden have been conducted under laboratory conditions and have not included inert surfaces in the community or food. Therefore, the objective of the study was to determine the presence of SARS-CoV-2 on food surfaces and other surfaces of public spaces in districts of Lima, Peru.

## Material and methods

### Study design

The study was observational, cross-sectional and descriptive.

### Population and sample

The population consisted of food surfaces and inert surfaces in the districts of San Martin de Porres, Villa el Salvador and San Juan de Lurigancho from Lima, Peru. The food samples were collected at local markets. The inert surfaces were collected at local markets, supermarkets, banking agencies and the system urban transport.

The districts of San Juan de Lurigancho, San Martin de Porres and Villa El Salvador, were selected because they represent the districts with the highest number of COVID-19 cases in Lima as of May 1^st^, 2020, according to the report of the National Center for Epidemiology, Prevention and Control of Diseases of the Ministry of Health.

In each district, the busiest markets, supermarkets and banking agencies were selected, as well as the public transport that had the longest travel through the districts. Non-probabilistic convenience sampling was applied to select the samples from food and inert surfaces. In the case of food, four samples of each food were collected for the determination of SARS-CoV-2 on the surface: two food samples and two control samples. The foods included were: bulk rice, avocado, banana, mango, lemon, tomato, lettuce, potato, cheese, chicken. For inert surfaces, five samples were taken for each commercial establishment (supermarkets, markets and banking agencies), considering another five control samples. In the case of inert surfaces in public transport, ten samples were taken per transport unit (three on railings and six on seats and one on the door). The frequency of sample collection was once a week and four times during a month from November to December 2020 as shown in table 1.

**Table 1.**
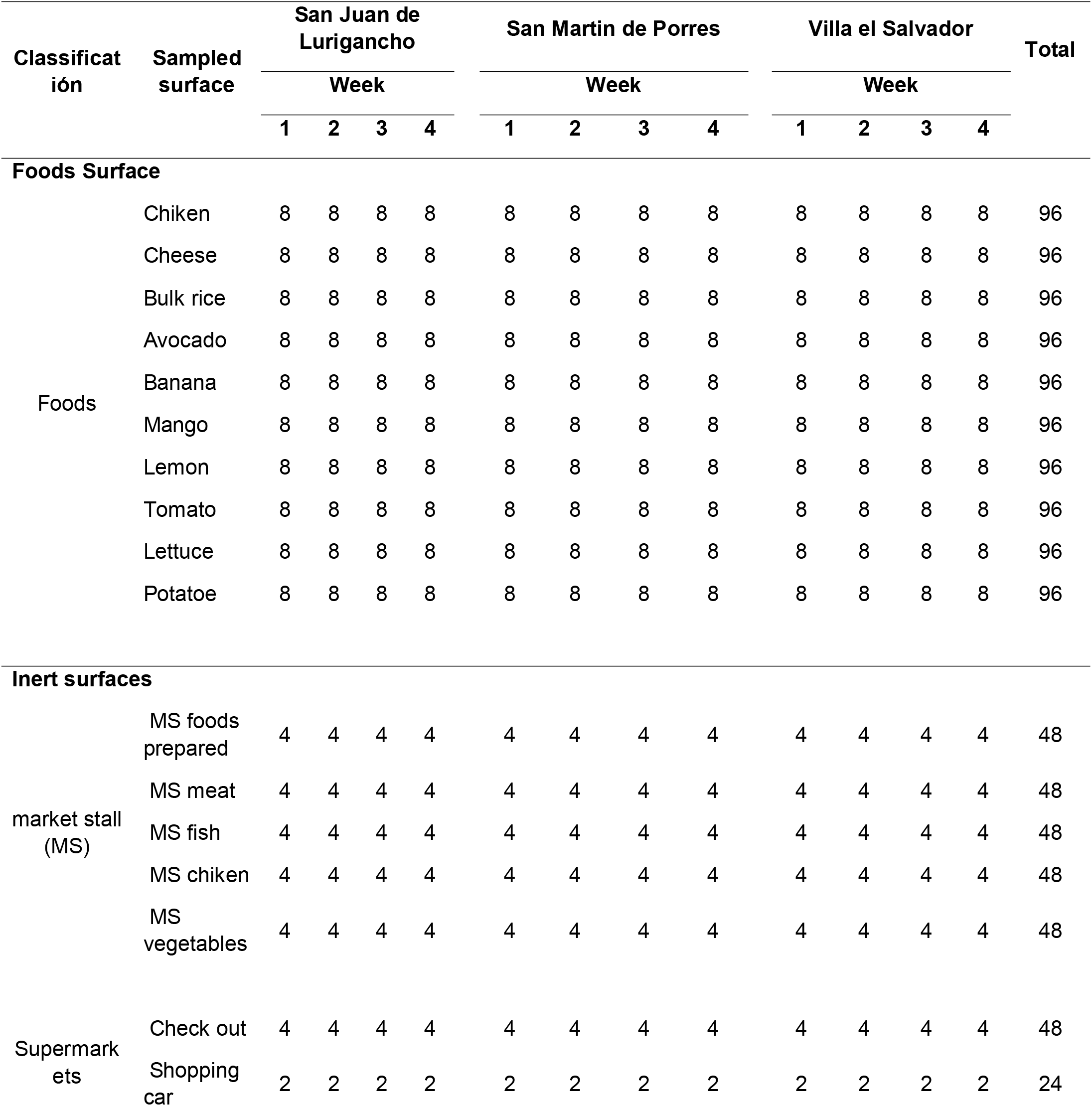
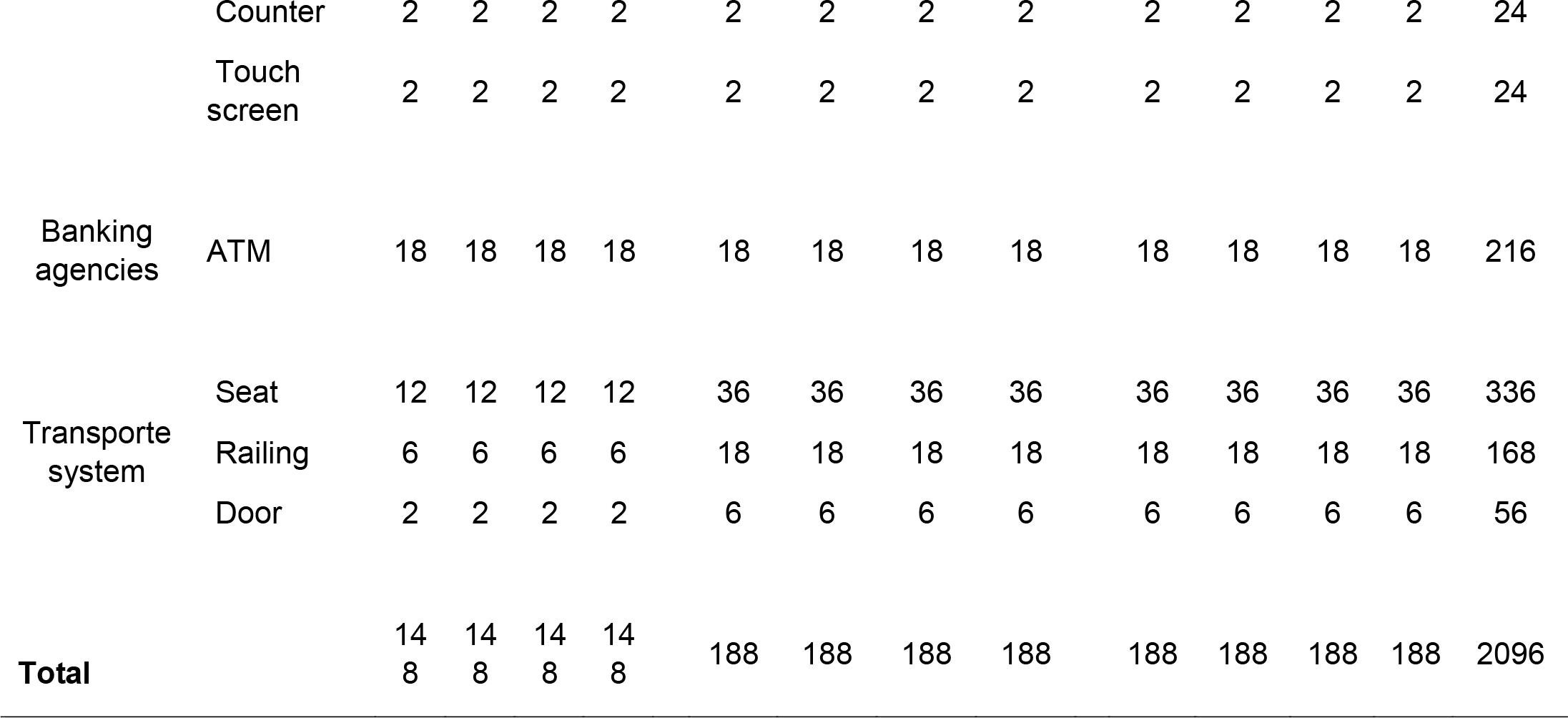
Quantity and type of surfaces sampled in 3 districts of Lima.

### Data collection

The data collection was carried out by technical laboratory personnel, trained in the collection, conservation and transport of the sample to the Peruvian national Institute of Health (INS). Prior to the collection of samples, authorization for the entry of field personnel was coordinated with those responsible for markets, supermarkets, banking agencies and transport units in each district. The data was collected during the months of November and December 2020. For the food samples, a regular purchasing procedure was carried out, so the technical staff went to the stalls, selected, bought the food and transported it to the laboratory. For inert surfaces, the technical staff in coordination with the person in charge of the commercial establishment indicated the surfaces to be sampled. The sample collection was carried out using the Jun Nuo Viral Transport Medium (VTM), the swabs were moistened in the VTM and passed on the food surfaces or inert surface. The VTMs were identified and kept in a cold chain during transportation to the National Reference Laboratory for Respiratory Viruses of the INS, where the samples were processed.

### Molecular diagnosis of SARS-CoV-2

The Zybio Nucleic Acid Detection Kit for SARS-CoV-2 - Magnetic Bead Method was used for the qualitative detection of RNA from swab samples. Briefly, the samples were homogenized and 200 µL were added to the 96-well extraction plate containing the Extraction Reagent (lysis buffer), then 15 µL of proteinase K was added and the plates were sealed with Parafilm. The RNA extraction was performed using the EMX-6000 (Zybio) equipment, where the Extraction Reagent I, Magnetic Bead Kit, Extraction Reagent II and Elution Reagent plates were placed following the instructions. The magnetic rack cover (blanket) was placed on the magnetic bead plate and 200 µL of nuclease-free water was added to the Extraction Reagent I plate (extraction control). For amplification, 5 µL of purified RNA were transferred to the amplification plate, from position A1 to E12 that includes 91 samples, the extraction control and aliquot control, 5 µL of the negative control and 5 µL were added positive control. The amplification plate was sealed and placed in the SANSURE MA6000 thermal cycler, using the fluorophores FAM (Gen *N*), ROX (Gen *ORF1ab*), VIC (Internal Control). According to the instructions of this qualitative kit, cycle threshold (Ct) greater than 39 for the ORF1ab gene are considered negative for the presence of molecular detection of the SARS-CoV-2 genetic material.

### SARS-CoV-2 viral isolation

The samples with a positive result of genetic material for SARS-CoV-2 were homogenized and filtered for sterile condition. For SARS-CoV-2 virus isolation the VERO 81 cell line (ATCC C1008) originated in African green monkey kidney was used. Vero-E6 cells were cultured in DMEM (Dulbecco’s Modified Eagles Medium) culture medium supplemented with streptomycin 100 mg/L, ampicillin 25 mg/L, 20% inactivated fetal bovine serum (SFB) and kept at 37 °C in a humid atmosphere of 5% CO2. The filtered sample was inoculated into the cells, and incubated for 7-10 days for the isolation of the SARS-CoV-2 virus. The presence of cytopathic effect was observed and the supernatant and cells were collected at the end of incubation. The diagnostic of isolation was determined by RT-PCR values.

### Study variables and indicators

For the purposes of the research objective, the variables analyzed were:

Presence of SARS-CoV-2: laboratory determination, the categories are presence or absence. Isolation: positive, negative; District: San Juan de Lurigancho (SJL), San Martin de Porres (SMP) and Villa El Salvador (VES). Food surface: bulk rice, avocado, banana, mango, lemon, tomato, lettuce, potato, cheese and chicken. Inert surface: handrails, seats, counter, touch screen, shopping cart handle, vending stand, and ATMs. Sample taking place: market, supermarket, public transport bus, train and bank agency.

### Analysis of data

The information registered in data collection sheets, as well as the results of the molecular tests were entered into an Excel database. The relative and absolute frequencies of the main variable, as well as of the secondary variables, were determined. A univariate analysis was performed, obtaining frequency distributions for the qualitative variables. The analysis was carried out in the statistical program SPSS V25.

### Ethical aspects

The study protocol was reviewed and approved by the INS Institutional Research Ethics Committee with the code OI-034-20.

## Results

2055 samples, 960 food surfaces and 1095 inert surfaces were collected and analyzed in 3 districts of Lima, Peru. No food surface sample was positive for the presence of SARS-CoV-2, while on inert surfaces the presence of the virus was only identified in one of the samples from ATMs by RT-PCR molecular test, from the San Martín de Porres district, which represented 1.4% of the samples collected in this type of surfaces (Table 02). The inner surface positive sample for SARS-CoV-2 have a Ct value of 35,51 for ORF1ab genes and no Ct for N gene.

**Table 02.**
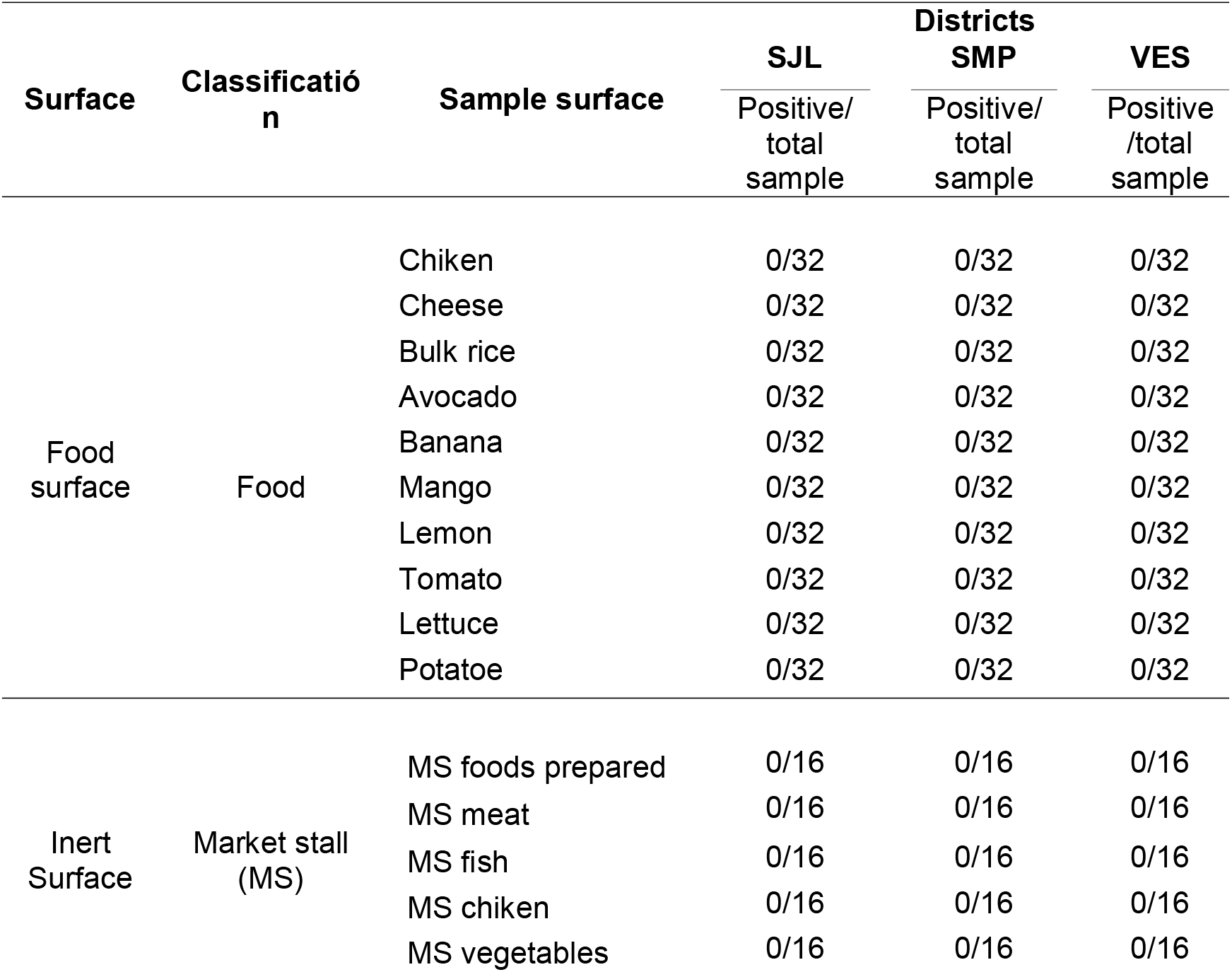
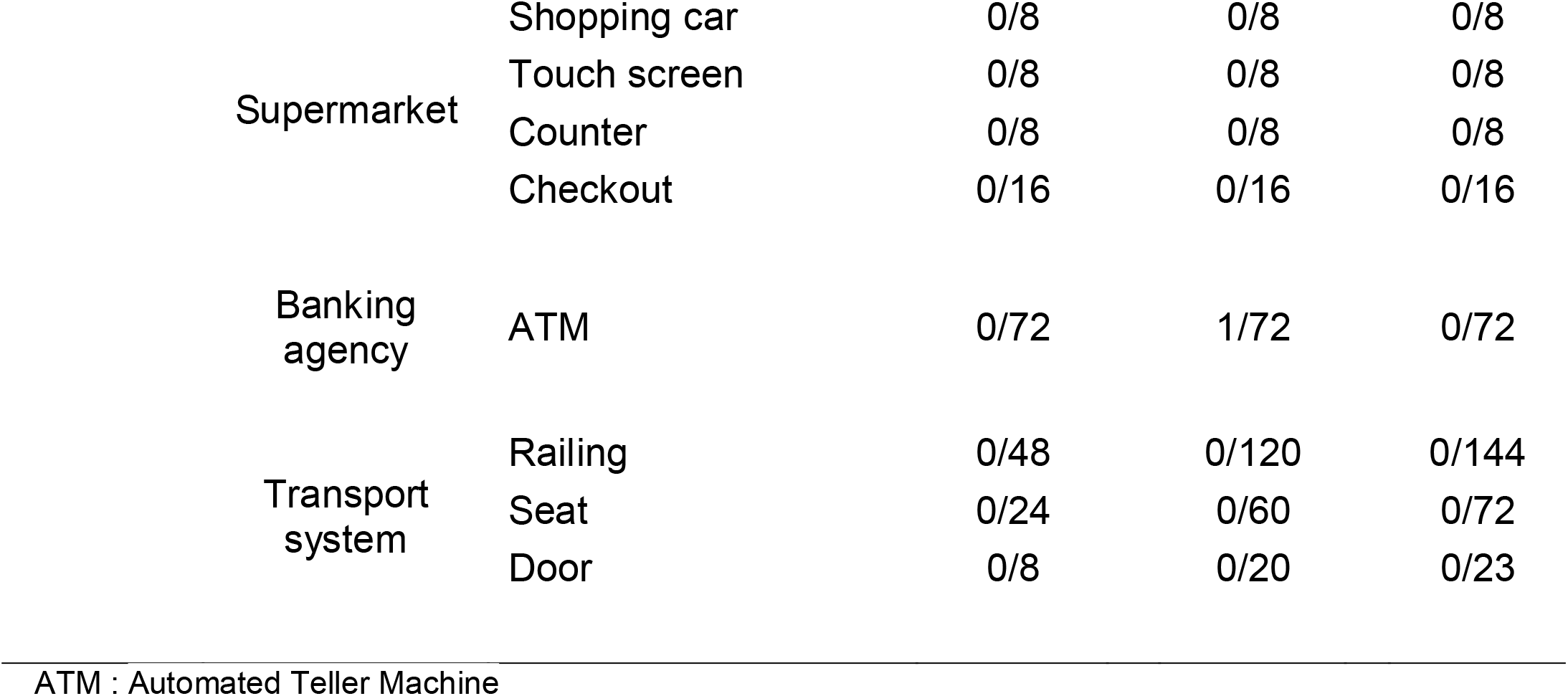
Results of the presence of SARS-CoV-2 (ARN) in food surfaces and inert surfaces of stalls, supermarkets, ATMs and transportation systems in 3 districts of Lima.

| Surface | Classificación | Sample surface | SJL | Districts SMP | VES |
| --- | --- | --- | --- | --- | --- |
|  |  |  | Positive/<br>total<br>sample | Positive/<br>total<br>sample | Positive<br>/total<br>sample |
| Food surface | Food | Chicken | 0/32 | 0/32 | 0/32 |
|  |  | Cheese | 0/32 | 0/32 | 0/32 |
|  |  | Bulk rice | 0/32 | 0/32 | 0/32 |
|  |  | Avocado | 0/32 | 0/32 | 0/32 |
|  |  | Banana | 0/32 | 0/32 | 0/32 |
|  |  | Mango | 0/32 | 0/32 | 0/32 |
|  |  | Lemon | 0/32 | 0/32 | 0/32 |
|  |  | Tomato | 0/32 | 0/32 | 0/32 |
|  |  | Lettuce | 0/32 | 0/32 | 0/32 |
|  |  | Potatoe | 0/32 | 0/32 | 0/32 |
| Inert Surface | Market stall (MS) | MS foods prepared | 0/16 | 0/16 | 0/16 |
|  |  | MS meat | 0/16 | 0/16 | 0/16 |
|  |  | MS fish | 0/16 | 0/16 | 0/16 |
|  |  | MS chicken | 0/16 | 0/16 | 0/16 |
|  |  | MS vegetables | 0/16 | 0/16 | 0/16 |

|  |  |  |  |  |
| --- | --- | --- | --- | --- |
| Supermarket | Shopping car | 0/8 | 0/8 | 0/8 |
|  | Touch screen | 0/8 | 0/8 | 0/8 |
|  | Counter | 0/8 | 0/8 | 0/8 |
|  | Checkout | 0/16 | 0/16 | 0/16 |
| Banking agency | ATM | 0/72 | 1/72 | 0/72 |
| Transport system | Railing | 0/48 | 0/120 | 0/144 |
|  | Seat | 0/24 | 0/60 | 0/72 |
|  | Door | 0/8 | 0/20 | 0/23 |

ATM : Automated Teller Machine

Subsequently, the SARS-CoV-2 positive sample was inoculated in Vero-81 cell culture for the virus isolation during 10 days. The supernatant and cells collected for virus isolation and the RT-PCR results showed a Ct value of 37,08 for ORF1ab gene and no Ct for N gene, indicating a a low viral load and not replicated virus, with probably RNA are remains of the inoculum of the original sample, additionally, no cytopatic effect was observed in the cuture. A subcultivation of virus isolation do not increase de viral charge, and finally was concluded as a negative sample for SARS-CoV-2. Ct values are showed in table 03.

**Table 03.**
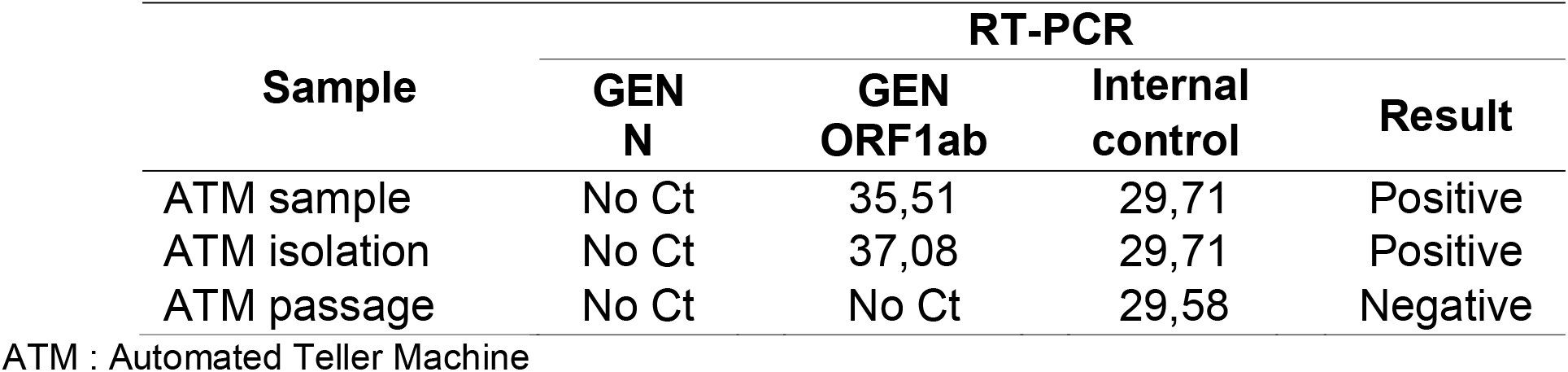
RT-PCR results for the ATM sample collected on an inert surface

| Sample | RT-PCR |  |  | Result |
| --- | --- | --- | --- | --- |
|  | GEN N | GEN ORF1ab | Internal control |  |
| ATM sample | No Ct | 35,51 | 29,71 | Positive |
| ATM isolation | No Ct | 37,08 | 29,71 | Positive |
| ATM passage | No Ct | No Ct | 29,58 | Negative |

ATM : Automated Teller Machine

## Discussion

The present study aimed to determine the presence of SARS-CoV-2 in food and inert surfaces of markets, supermarkets, banks and means of transportation in 3 districts of Lima, Peru. The results show that only the presence of SARS-CoV-2 was found on the inert surface of one ATM machine, while on other surfaces, including food, the presence of this virus was not identified.

As far as we investigated, this is one of the first reports of the presence and stability of SARS-CoV-2 in fresh food (without refrigeration) and in real conditions (outside the laboratory), therefore, the negative results of the presence of SARS-CoV-2 cannot be directly compared with the available literature. Yepiz-Goméz et al studied the recovery and survival efficiency of two respiratory viruses, an adenovirus (HAdV-2) and a coronavirus other than SARS-CoV-2 (HCoV-229E) in lettuce, strawberries and raspberries stored at 4 °C (refrigeration) and reported that the coronavirus was able to recover in the first 3 days with an efficiency of up to 19.6% (16). Meanwhile, Dai et al, evaluated the survival of SARS-CoV-2 in salmon, which was artificially infected with SARS-CoV-2 inoculum, dried with filter paper and stored at room temperature (25 °C) and refrigeration (4 °C) and found that the meat preserved at room temperature showed viable virus up to two days, while the refrigerated sample persisted until 8 days (17).

The reasons why we did not find the presence of SARS-CoV-2 in the analyzed food samples could be due, among others, to the fact that the samples were collected during the period of greatest fall in COVID-19 infections in Peru. Second, the foods that were purchased in markets and supermarkets could have been exposed to cleaning substances such as alcohol, bleach, etc, which could reduce the number of viral particles. In addition, unlike the studies in food that report the presence of SARS-CoV-2, in the present study the food was not artificially infected. We aimed to find contaminated food by the manipulation and interaction of people in the markets. Also, the food was kept in the markets and transported to the laboratory at room temperature (27 °C). The negative results for the presence of SARS-CoV-2 on fresh food surfaces are consistent with the EFSA and Food and Drug Agency recommendations that there is no evidence of transmission of COVID-19 through food (14,18). Likewise, the International Commission on Microbiological Specifications for Foods has prepared a joint declaration of the low risk of transmission of SARS-CoV-2 by food (19).

Additionally, in the case of inert surfaces, the presence of SARS-CoV-2 RNA was only found in one sample (0.06%). In this sense, the evidence is consistent with that of previous studies that used the same methodology on inert surfaces of hospital wards (20,21,22), in which no positive results were found. On the other hand, other studies evaluated the presence of SARS-CoV-2 genetic material on other high-traffic surfaces such as buses and trains (23); handles for garbage cans, or for entry and exit of essential shops (24); or parks and water sources (25). These studies did find positive results, which ranged from 4-8%.

The relative contradiction between these results can be explained by some factors; For example, Kwon et al (2021) postulate that environmental temperature plays an important role in the stability of this virus, making it difficult to detect as temperature increases, such as in summer (26). In our study, the collection of the samples took place between the months of November to December, in which the temperatures in the city of Lima oscillate on average between 20 ° – 22 ° C, which could explain the absence of positive results. On the other hand, the material in which the virus is found can influence its detection; being its average duration in plastic and steel between three to four days (11). These are the materials from which most of the inert surfaces evaluated in this study are made.

The strengths of the study carried out are a robust sampling design that covers the geographical and temporal dimension, which allowed the collection of samples in 3 densely populated districts, during 4 consecutive weeks and 4 repetitions in each sample of 10 types of food and 13 types of inert surfaces, making a total of 2016 samples. Likewise, the transfer of the samples was carried out in means of viral transport and fulfilling the technical criteria; the collection of the samples in the field and the analysis in the laboratory was carried out by personnel trained and experienced in respiratory viruses. In addition, unlike previous studies carried out in the laboratory using spiked samples, our study was carried out in real conditions trying to demonstrate alternative routes of infection, rather than microdrops between people.

The study also has limitations, such as that the collection of samples was carried out in the months of November and December, a period in which the number of new cases was less than 1,000 in Lima, Peru, according to the National Center for Epidemiology, Prevention and Control of Diseases. It is important to indicate that the World Health Organization mentions that the biological basis for the possibility of infection through contact with surfaces and food is related to fomites, which is still considered a concept with contradictory evidence (27). Furthermore, the study was carried out only in districts of Lima, Peru, and not in other regions of the country; however, it should be considered that these districts were the ones that showed the highest number of infections in the most critical period of the pandemic in Peru.

Another limitation, is the lower isolation of virus from samples with higher Ct value, Ct > 31 by RT-PCR method. The rt-PCR Ct value correlate strongly with cultivable virus and likelihood of infectiosness, In this sense, a higher number of positive samples, will offer us the probability of a greater number of virus isolation (11). An additional aspect to consider is that, in the study period, frequent cleaning of the surfaces to be studied was observed, which could have had an impact triggering lower possibilities of obtaining positive results, but not on the elimination of their detection as has been described in previous experiences (23).

Peru has been one of the most affected countries by the COVID-19 pandemic and although it adopted social immobilization measures early, some essential activities were maintained such as the provision of food in markets, supermarkets, banking services and means of transport, places where conditions could be generated for the direct and indirect contagion of SARS-CoV-2. The results of the study show the presence of SARS-CoV-2 only on the surface of ATMs, which is consistent with the current literature that reports specific findings of this virus on various surfaces. It should be noted that the absence of detection of SARS-CoV-2 in most surfaces does not indicate at all to suspend measures such as disinfection of surfaces and food, since these public health strategies not only increase the probability of reducing the viral transmission (whatever the magnitude) (27); but, more importantly, they help to cut the chain of transmission of other pathogens, such as enteropathogenic bacteria, which also represent a health problem in Latin American health systems (22).

In conclusion, the results of the study show that no presence of SARS-CoV-2 was found on food surfaces at room temperature such as cereals, fruits, dairy vegetables and meats, although these foods could have received some cleaning treatment before their analysis. Regarding inert surfaces, the presence of not infective SARS-CoV-2 virus was identified in only one sample corresponding to ATM, while the presence of SARS-CoV-2 was not verified on other inert surfaces. Despite the negative results, the frequency of disinfection measures should be maintained and increased, especially on inert high-contact surfaces, and hygiene measures on food should continue. It is necessary to develop new studies in periods of high contagion in the community, include refrigerated and frozen foods and take into account the new variants of COVID-19 that are circulating in our country.

## Data Availability

All data produced in the present study are available upon reasonable request to the authors

## CONTRIBUTIONS

Conceptualization: LH, KA, JPA, EG, YA, HM, DF. Data collect EG, PH, YA, HM, DF. Laboratory sample analysis: OE, MP. Writing – original draft: JPA, YA, KA, HM, OE, MP. Writing – review & editing: KA, YA, OE, DF, MP, EG, HM, PH, LH, JPA.

## Bibliography

1. Rothan HA, Byrareddy SN. The epidemiology and pathogenesis of coronavirus disease (COVID-19) outbreak. J Autoimmun. 2020;109: 102433. https://doi.org/10.1016/j.jaut.2020.102433 Epub 2020 Feb 26. PMID: 32113704; PMCID: PMC7127067.

2. World Health Organization. Situation report - 135 [Internet]. OMS; 2020 [citado 4 de junio de 2020]. Disponible en: https://www.who.int/emergencies/diseases/novel-coronavirus-2019/situation-reports

3. Ahn DG, Shin HJ, Kim MH, Lee S, Kim HS, Myoung J, et al. Current Status of Epidemiology, Diagnosis, Therapeutics, and Vaccines for Novel Coronavirus Disease 2019 (COVID-19). J Microbiol Biotechnol. 2020; 30(3): 313–324. https://doi.org/10.4014/jmb.2003.03011 PMID: 32238757.

4. Gobierno del Perú. Presidente Vizcarra dio a conocer primer caso de infección por coronavirus en el Perú e hizo un llamado a la población a mantener la calma [Internet]. 2020 [citado 5 de abril de 2020]. Disponible en: https://www.gob.pe/institucion/presidencia/noticias/86976-presidente-vizcarra-dio-a-conocer-primer-caso-de-infeccion-por-coronavirus-en-el-peru-e-hizo-un-llamado-a-la-poblacion-a-mantener-la-calma

5. Ministerio de Salud. Minsa lamenta el sensible fallecimiento de dos personas por infección por COVID-19 (Comunicado N°21) [Internet]. 2020 [citado 5 de abril de 2020]. Disponible en: https://www.gob.pe/institucion/minsa/noticias/109603-minsa-lamenta-el-sensible-fallecimiento-de-dos-personas-por-infeccion-por-covid-19-comunicado-n-21

6. Presidencia del Consejo de Ministros. Decreto Supremo N° 094-2020-PCM. Medidas para la ciudadanÍa hacia una nueva convivencia y prórroga del Estado de Emergencia [Internet]. may 23, 2020. Disponible en: https://www.gob.pe/institucion/pcm/normas-legales/584231-094-2020-pcm

7. Liu Y, Gayle AA, Wilder-Smith A, Rocklöv J. The reproductive number of COVID-19 is higher compared to SARS coronavirus. J Travel Med [Internet]. 13 de marzo de 2020 [citado 5 de abril de 2020];27(2). Disponible en: https://academic.oup.com/jtm/article/27/2/taaa021/5735319

8. Riou J, Althaus CL. Pattern of early human-to-human transmission of Wuhan 2019 novel coronavirus (2019-nCoV), December 2019 to January 2020. Euro surveill. 2020; 25(4):2000058. https://doi.org/10.2807/1560-7917.ES.2020.25.4.2000058

9. Organización Mundial de la Salud. Modes of transmission of virus causing COVID-19: implications for IPC precaution recommendations [Internet]. 2020 [citado 4 de junio de 2020]. Disponible en: https://www.who.int/publications-detail/modes-of-transmission-of-virus-causing-covid-19-implications-for-ipc-precaution-recommendations

10. Fathizadeh H, Maroufi P, Momen-Heravi M, Dao S, Köse Ş, Ganbarov K, et al. Protection and disinfection policies against SARS-CoV-2 (COVID-19). Infez Med. 2020;28(2):185–191. PMID: 32275260. http://www.ncbi.nlm.nih.gov/pubmed/32275260

11. Van Doremalen N, Bushmaker T, Morris DH, Holbrook MG, Gamble A, Williamson BN, et al. Aerosol and Surface Stability of SARS-CoV-2 as Compared with SARS-CoV-1. N Engl J Med. 2020; 382(16):1564–1567. https://doi.org/10.1056/nejmc2004973 Epub 2020 Mar 17. PMID: 32182409; PMCID: PMC7121658.

12. Kampf G, Todt D, Pfaender S, Steinmann E. Persistence of coronaviruses on inanimate surfaces and their inactivation with biocidal agents. J Hosp Infect. marzo de 2020;104(3):246–251. https://doi.org/10.1016/j.jhin.2020.01.022 Epub 2020 Feb 6. Erratum in: J Hosp Infect. 2020 Jun 17;: PMID: 32035997; PMCID: PMC7132493.

13. Ong SWX, Tan YK, Chia PY, Lee TH, Ng OT, Wong MSY, et al. Air, Surface Environmental, and Personal Protective Equipment Contamination by Severe Acute Respiratory Syndrome Coronavirus 2 (SARS-CoV-2) From a Symptomatic Patient. JAMA. 2020; 323(16):1610–1612. https://doi.org/10.1001/jama.2020.3227 PMID: 32129805; PMCID: PMC7057172.

14. European Food Safety Authority. Coronavirus: no evidence that food is a source or transmission route [Internet]. European Food Safety Authority. 2020 [citado 21 de abril de 2020]. Disponible en: https://www.efsa.europa.eu/en/news/coronavirus-no-evidence-food-source-or-transmission-route

15. Van Kerkhove MD. Brief literature review for the WHO global influenza research agenda--highly pathogenic avian influenza H5N1 risk in humans. Influenza Other Respir Viruses. 2013; 2:26–33. https://doi.org/10.1111/irv.12077 PMID: 24034480.

16. Yépiz-Gómez MS, Gerba CP, Bright KR. Survival of Respiratory Viruses on Fresh Produce. Food Environ Virol. 2013 17;5(3):150–6. https://doi.org/10.1007/s12560-013-9114-4 Epub ahead of print. PMID: 23681671; PMCID: PMC7091382.

17. Dai M, Li H, Yan N, Huang J, Zhao L, Xu S, Wu J, Jiang S, Pan C, Liao M. Long-term Survival of SARS-CoV-2 on Salmon as a Source for International Transmission. J Infect Dis. 2021; 223(3): 537–539. https://doi.org/10.1093/infdis/jiaa712 PMID: 33179031; PMCID: PMC7717293.

18. Shea K, WoodCoock J, Actualización sobre el COVID-19: El USDA y la FDA enfatizan que la información epidemiológica y cientÍfica actual indica que no hay transmisión del COVID-19 a través de los alimentos o de los envases de los alimentos. Press Announcements. [Internet]. Food and Drug Agency. 2021. [citado 11 de agosto de 2021]. Disponible en: https://www.fda.gov/news-events/press-announcements/actualizacion-sobre-el-covid-19-el-usda-y-la-fda-enfatizan-que-la-informacion-epidemiologica-y

19. International Commission on Microbiological Specifications for Foods. ICMSF Opinion on SARS-CoV-2 and its relationships to food safety. [Internet] ICMSF 2020.. [citado 11 de agosto de 2021]. Disponible en: https://www.icmsf.org/wp-content/uploads/2020/09/ICMSF2020-Letterhead-COVID-19-opinion-final-03-Sept-2020.BF_.pdf

20. Wang J, Feng H, Zhang S, Ni Z, Ni L, Chen Y, Zhuo L, Zhong Z, Qu T. SARS-CoV-2 RNA detection of hospital isolation wards hygiene monitoring during the Coronavirus Disease 2019 outbreak in a Chinese hospital. International Journal of Infectious Diseases. 2020; 94: 103–106. ISSN 1201-9712. https://doi.org/10.1016/j.ijid.2020.04.024

21. Colaneri M, Seminari E, Piralla A, Zuccaro V, Filippo AD, Baldanti F, Bruno R, Mondelli MU; COVID19 IRCCS San Matteo Pavia Task Force. Lack of SARS-CoV-2 RNA environmental contamination in a tertiary referral hospital for infectious diseases in Northern Italy. J Hosp Infect. 2020;105(3): 474–6. https://doi.org/10.1016/j.jhin.2020.03.018 Epub ahead of print. PMID: 32201338; PMCID: PMC7156210.

22. Gavaldà-Mestre L, RamÍrez-Tarruella D, Gutiérrez-Milla C, Guillamet-Roig F, Orriols-Ramos R, Tisner SR, Pàrraga-Niño N. Nondetection of SARS-CoV-2 on high-touch surfaces of public areas next to COVID-19 hospitalization units. Am J Infect Control. 2021: S0196-6553(21)00007-9. https://doi.org/10.1016/j.ajic.2021.01.007 Epub ahead of print. PMID: 33450309; PMCID: PMC7837185.

23. Moreno T, Pintó RM, Bosch A, Moreno N, Alastuey A, Minguillón MC, Anfruns-Estrada E, Guix S, Fuentes C, Buonanno G, Stabile L, Morawska L, Querol X. Tracing surface and airborne SARS-CoV-2 RNA inside public buses and subway trains. Environ Int. 2021; 147, 106326. ISSN 0160-4120. https://doi.org/10.1016/j.envint.2020.106326

24. Harvey AP, Fuhrmeister ER, Cantrell M, Pitol AK, Swarthout JM, Powers JE, Nadimpalli ML, Julian TR, Pickering AJ. Longitudinal monitoring of SARS-CoV-2 RNA on high-touch surfaces in a community setting. medRxiv [Preprint]. 2020 Nov 1:2020.10.27.20220905. https://dx.doi.org/10.1101%2F2020.10.27.20220905.20220905 PMID: 33140065; PMCID: PMC7605577.

25. Kozer E, Rinott E, Kozer G, Bar-Haim A, Benveniste-Levkovitz P, Klainer H, Perl S, Youngster I. Presence of SARS-CoV-2 RNA on playground surfaces and water fountains. Epidemiol Infect. 2021;149:e67. https://doi.org/10.1017/S0950268821000546 PMID: 33678202; PMCID: PMC7985893.

26. Kwon T, Gaudreault NN, Richt JA. Environmental Stability of SARS-CoV-2 on Different Types of Surfaces under Indoor and Seasonal Climate Conditions. Pathogens. 2021;10(2):227. https://doi.org/10.3390/pathogens10020227 PMID: 33670736; PMCID: PMC7922895.

27. Marquès M, Domingo JL. Contamination of inert surfaces by SARS-CoV-2: Persistence, stability and infectivity. A review. Environ Res. 2021 Feb;193:110559. https://doi.org/10.1016/j.envres.2020.110559 Epub 2020. PMID: 33275925; PMCID: PMC7706414.

